# Are there causal associations between obsessive-compulsive disorder and cardiometabolic phenotypes? A genetic correlation and bi-directional Mendelian randomization study

**DOI:** 10.1101/2025.04.08.25325472

**Authors:** Robyn E. Wootton, James J. Crowley, Josep Pol-Fuster, Anna Holmberg, Christian Rück, Psychiatry Genomics Consortium–Obsessive-Compulsive Disorder Working Group, David Mataix-Cols, Lorena Fernández de la Cruz

## Abstract

In epidemiological studies, obsessive-compulsive disorder (OCD) is robustly associated with increased risk of cardiometabolic disorders, including cardiovascular diseases, type 2 diabetes, and obesity. However, the mechanisms behind these associations are unclear. We conducted genetic correlation analyses to explore shared genetic etiology and bi-directional summary-level Mendelian randomization (MR) to explore potential causal effects between genetic liability to OCD and 14 cardiometabolic phenotypes (e.g., coronary artery disease, blood pressure, body mass index [BMI]). If causal effects were observed, we planned to conduct multivariable-MR to explore indirect effects via health behaviors. We found no evidence for genetic correlations between OCD and any of the cardiometabolic phenotypes under study, except for a negative correlation with BMI (rG=-0.123, SE=0.029, p<0.001). Summary-level MR showed no evidence for causal effects. Therefore, multivariable-MR was not conducted. We found limited evidence for shared genetic etiology or causal effects. However, we were only powered to detect medium effects in the direction of OCD to cardiometabolic traits, leaving the possibility of smaller causal effects existing. Future studies with larger, more representative samples will help to further interpret findings.

## INTRODUCTION

Obsessive-compulsive disorder (OCD) is a chronic mental disorder affecting around 1-2% of the population (1). Epidemiological studies have shown a robust link between OCD and morbidity and mortality due to endocrine, metabolic, and circulatory system disorders, such as cardiovascular diseases (CVD), type 2 diabetes, and obesity (2-5). However, the mechanisms behind these associations are unclear.

Population-based studies using quasi-experimental discordant sibling designs have shown that the associations between OCD and cardiometabolic disorders survive adjustment for unmeasured familial confounding (2-4). Consistent with these findings, familial coaggregation analyses showed limited evidence for shared familial risk between OCD and CVD, obesity, type 2 diabetes, and hyperlipidemia (6). Taken together, these epidemiological results seem to suggest that the contribution of shared genetic risk factors to the association between OCD and cardiometabolic disorders is small or negligible.

Along the same lines, the largest OCD genome-wide association study (GWAS) to date, including over 50,000 cases and 30 OCD-associated loci, did not find significant genetic correlations between OCD and several cardiovascular and metabolic phenotypes, including myocardial infarction, coronary artery disease, type 2 diabetes, cholesterol, and triglycerides (7). However, the same study found significant negative correlations with hip circumference, body fat, and body mass index (BMI). Thus, the pattern of genetic correlations may differ according to the specific phenotype under study. Moreover, although causal effects are more likely when genetic correlations are identified, the absence of genetic correlation does not preclude the presence of a causal effect (8).

A complementary method to further understand the nature of the association between OCD and cardiometabolic outcomes is Mendelian randomization (MR). MR is a genetically informed design for potential causal interference that treats genetic variation as a natural experiment in which individuals are genetically predisposed to higher versus lower mean levels of an exposure during their lifetime (9). Because genetic variants are considered to be allocated randomly before birth, they are relatively independent of environmental factors and established well before onset of disease, minimizing the issues with residual confounding and reverse causation that so often appear in observational studies (9, 10). To our knowledge, three MR studies have explored the causal associations between OCD and cardiometabolic outcomes. One study found that OCD was not causally associated with CVD (11), while the other two found evidence of a causal negative association between obesity (operationalized as higher BMI or waist-hip ratio) and OCD (12, 13). These studies focused on one single cardiometabolic phenotype and were limited by the use of older OCD GWAS summary statistics (e.g., 14). With the recent publication of the largest OCD GWAS to date (7), and greater variance explained by the genetic instrument, we now have improved power to assess causal effects between OCD and cardiometabolic phenotypes.

This study investigated the potentially causal associations between OCD and 14 cardiometabolic disorders and traits by computing genetic correlations and bi-directional MR. We used the latest summary statistics from the first adequately powered OCD GWAS (7) and from a broad range of cardiometabolic phenotypes with publicly available GWAS. Our specific aims were to: 1) compute genetic correlations to determine the genetic overlap between OCD and cardiometabolic disorders; 2) perform bi-directional univariable MR to test if liability to OCD causally increases cardiometabolic risk, and vice versa; and 3) perform multivariable MR to test if key behaviors (e.g., alcohol consumption, use of medication) mediate effects of liability to OCD on cardiometabolic risk, if causal associations were found.

## METHODS

### Data sources and measures

#### Obsessive-compulsive disorder

The most recent GWAS of OCD comprised 53,660 cases and 2,044,417 controls (7). Cases were identified either through medical records, clinician assessment or self-report. There were a total of 30 independent genome-wide significant single nucleoid polymorphisms (SNPs) identified, and the total SNP-based heritability was 6.7%.

#### Cardiometabolic phenotypes

The included 14 cardiometabolic traits were: coronary artery disease, myocardial infarction, heart failure, heart rate variability, triglycerides, total cholesterol, high-density lipoprotein (HDL) cholesterol, low-density lipoprotein (LDL) cholesterol, type 2 diabetes, body mass index (BMI), ischemic stroke, systolic blood pressure, diastolic blood pressure, and thrombosis.

#### Positive control

As this was the first study to apply MR to OCD as a genetic instrument, we included suicide attempts as a positive control outcome, where we hypothesized a causal effect of genetic liability to OCD on increased risk of suicide attempts (15-17). To this end, we used the most recent available GWAS of suicide attempts, comprising 35,786 cases and 779,392 controls of European ancestry (18). As cardiometabolic traits have previously been used as MR instruments and had greater power, we did not employ positive controls in the opposite direction.

#### GWAS summary statistics

All analyses were conducted using publicly available summary statistics from previously conducted GWAS, summarized in **Table 1**. We prioritized the largest available GWAS of each trait with the least possible sample overlap (as this can bias results towards the confounded estimate when instruments are weak) (19), and conducted in individuals of European ancestry—with the exception of myocardial infarction, where available summary statistics were of mixed ancestry (20), so we performed a replication using FinnGen data (21) (**Supplementary Note 1**). SNP effects for all continuous traits were converted to standardized betas and SNP effects for binary traits were log odds ratios.

### Statistical analysis

#### Genetic correlations

Genetic correlations were estimated between OCD and each of the cardiometabolic traits using cross-trait LD Score Regression v1.0.1 (22). This method estimates the degree of concordance between effect sizes for the same SNPs across the two traits, while additionally accounting for the degree of linkage disequilibrium (LD) between SNPs. A SNP’s effect size will be inflated, as this effect includes the effects of the SNPs in LD. Therefore, if a trait is polygenic, SNPs with higher LD will show larger effect sizes than those with lower LD. The GWAS estimate for a particular SNP incorporates the effects of all other SNPs that are in LD with that SNP. This information is used to compute an overall genetic correlation between two phenotypes across all SNPs (22).

#### Mendelian randomization analysis using summary-level data

Summary level (or two-sample) MR was applied to explore evidence for bi-directional causal effects between liability to OCD and a range of cardiometabolic traits. MR utilizes genetic variants as instrumental variables to estimate the causal effect of an exposure (e.g., OCD) on an outcome (e.g., cardiometabolic phenotypes). Three core assumptions must be satisfied in order to infer causality: (1) the relevance assumption—the genetic instrument must be robustly associated with the exposure; (2) the independence assumption—there must be no confounders of the genetic instrument and the outcome; and (3) the exclusion-restriction assumption—the genetic instrument must only be associated with the outcome via the exposure (23). The exclusion-restriction assumption can be violated in the presence of horizontal pleiotropy, which occurs when the genetic instruments for the exposure are also associated with confounders or the outcome via other pathways. We use a range of sensitivity analyses (outlined below) to explore the likelihood of assumption violations and bias due to pleiotropy.

All analyses were conducted using the TwoSampleMR package, version v0.4.1732 for R. We used four different MR methods: inverse-variance weighted, MR Egger, weighted median, and weighted mode. Each method makes different assumptions about the presence of pleiotropy and therefore a consistent effect across multiple methods strengthens causal evidence. If a SNP was unavailable in the outcome GWAS summary statistics, then proxy SNPs were searched for with a minimum linkage disequilibrium r2 = 0.8, and palindromic SNPs were aligned if minor allele frequency (MAF) < 0.3. We estimated instrument strength by calculating the mean F-statistic across all SNPs, where an F-statistic < 10 is considered weak. We calculated the regression dilution I^2^_GX_ as an indicator of the suitability of the instrument for MR Egger. When I^2^_GX_ was between 0.6 and 0.9, a SIMEX correction was performed. We performed Rucker’s Q test of heterogeneity and the MR Egger intercept test to estimate potential directional horizontal pleiotropy. We performed Steiger filtering to check that all genetic variants explained more variance in the exposure than the outcome. If this were not the case, it could suggest potential reverse causation. When there was evidence for causal effects, scatter plots and leave-one-out SNP plots were visually inspected to check for possible outliers.

Where exposures were binary, effect estimates were multiplied by 0.693 so that units can be interpreted as a change per doubling in the odds of the exposure. When outcomes were binary, effect estimates are presented as odds ratios. When outcomes were continuous, effect estimates are presented as standardized betas.

We estimated the minimum effect size for 80% power in all univariable MR analyses using the mRnd power calculator (24). Power calculations did not guide the analyses conducted but were used in effect interpretation.

#### Multivariable Mendelian randomization

If evidence for causal effects were observed in MR analyses, we planned to follow up using summary-level multivariable MR (MVMR) (25). MVMR is an extension of the MR method where multiple exposures can be included to explore possible indirect pathways. Estimates of one exposure, after conditioning on the other exposure can reveal whether effects are independent. Where we found evidence for causal effects in univariable MR of liability to OCD on cardiometabolic traits, we planned to investigate whether direct effects were explained through health behaviors: alcohol consumption (26), tobacco smoking (26, 27), physical activity (28), BMI (29), and diet (30). We planned to conduct analyses using the ‘MVMR’ package in R (31) using genome-wide summary statistics (a two-sample approach). We planned to estimate F-statistics conditional on the other exposure to ensure instruments were of sufficient strength (31).

## RESULTS

### Genetic correlations

The results of genetic correlations between OCD and cardiometabolic phenotypes are shown in **Figure 1**. As anticipated, there was evidence for a positive genetic correlation between OCD and our positive control phenotype of suicide attempts (rG = 0.365, SE = 0.058, p<0.001). There was no evidence for a significant overall genetic correlation between OCD and any of the cardiometabolic traits, with the exception of BMI, which was significantly negatively genetically correlated (rG = -0.123, SE = 0.029, p<0.001).

### Mendelian randomization

#### Power analyses and instrument strength

When OCD was the exposure and cardiometabolic phenotypes were the outcome, power calculations demonstrated we only had 80% power to detect medium effect sizes in MR analyses, with the exception of power to detect small effects on ischemic stroke (**Supplementary Table 1**). In the reverse direction, we had 80% power to detect small effects on OCD when cardiometabolic traits were the exposure, with the exception of ischemic stroke and heart failure, where we only had power to detect medium effects (**Supplementary Table 1**).

All genetic instruments had sufficient instrument strength to conduct MR analyses with reduced risk of weak instrument bias, as evidenced by a mean F statistic > 10 (**Supplementary Table 2**).

#### Effects of OCD on cardiometabolic outcomes

The primary MR method was the inverse-variance weighted method, which assumes balanced horizontal pleiotropy. It was not possible to conduct MR Egger sensitivity test when OCD was the exposure due to low regression dilution (**Supplementary Table 2**). Therefore, only weighted median and weighted mode were used as sensitivity methods.

As anticipated, there was evidence to support a causal effect of liability to OCD on our positive control outcome of suicide attempts (**Figure 2**) such that per doubling in the odds of OCD, there was a 23% increased odds of suicide attempts (95% CI: 4% – 46%, p=0.001). Effect estimates were highly consistent across the sensitivity methods weighted median and weighted mode (**Supplementary Figures 1 and 2**). Visual inspection did not suggest results were biased by outlying SNPs (**Supplementary Figures 4 and 5**).

MR analyses did not provide evidence to support a causal effect of OCD on any of the cardiometabolic traits (**Figure 2**), and this was consistent across weighted median and weighted mode sensitivity methods (**Supplementary Figures 1 and 2**). There was evidence for significant heterogeneity for several outcomes (thrombosis, BMI, HDL cholesterol, total cholesterol, triglycerides, and blood pressure) (**Supplementary Table 3**), which could be the result of pleiotropy. However, the MR Egger intercepts suggest that there is significant bias from directional horizontal pleiotropy (with the exception of total cholesterol, triglycerides, and diastolic blood pressure) (**Supplementary Table 4**). There was limited evidence from Steiger filtering for bias from reverse causation (**Supplementary Table 5**), with > 90% of genetic variants explaining more variance in the exposure than the outcome for all traits except type 2 diabetes, where 50% of the variants explained more variance in the outcome.

#### Effects of cardiometabolic traits on OCD risk

When cardiometabolic traits were the exposures, there was no evidence of regression dilution bias, with the exception of ischemic stroke (**Supplementary Table 2**). Therefore, MR Egger could be employed as a sensitivity test in the majority of analyses, alongside weighted median and weighted mode.

MR analyses did not provide evidence to support a causal effect of any cardiometabolic traits on odds for OCD (**Figure 3**), and this was consistent across the sensitivity methods (**Supplementary Figure 3**). There was no evidence for significant heterogeneity across all traits (**Supplementary Table 3**), further supported by the MR Egger intercept tests, which suggested no bias from directional horizontal pleiotropy (**Supplementary Table 4**). Furthermore, Steiger filtering found no evidence for bias from reverse causation (**Supplementary Table 5**), with 100% of genetic variants explaining more variance in the exposure than the outcome.

### Multivariable Mendelian randomization

We did not find strong evidence for causal effects of liability to OCD on cardiometabolic outcomes in univariable MR, and therefore MVMR follow-up analyses were not conducted to explore possible mechanisms.

## DISCUSSION

At the population level, OCD has been shown to be robustly associated with a range of cardiometabolic disorders (3-5). The current study systematically investigated whether these associations could be due to shared genetic etiology by conducting genetic correlations and due to causal effects by conducting bi-directional MR analyses between OCD and 14 cardiovascular and metabolic phenotypes. The results showed that there was no genetic correlation between OCD and any of the cardiometabolic traits, except for BMI, which showed a negative correlation with OCD. Additionally, MR results showed a lack of evidence for causal effects in either direction.

The lack of genetic correlations across a range of cardiometabolic phenotypes replicates the results of the genetic correlations reported in the latest OCD GWAS (7), showing no correlation between OCD and the whole cluster of 8 cardiovascular phenotypes. Therefore, the observed associations between OCD and cardiometabolic outcomes are unlikely due to shared genetic etiology (at least not from common variants). This is consistent with evidence from previous epidemiological studies using family-based designs, including sibling (3, 4) and familial coaggregation designs (6). After controlling for shared genetic and environmental factors, these population-based results suggested that the associations between OCD and cardiometabolic outcomes were largely independent from familial factors, suggesting a possible causal effect. However, our MR analyses showed a lack of evidence to support causal effects in either direction. This could be the result of low statistical power, as we only had power for medium effects in the direction of OCD to cardiometabolic phenotypes and previous evidence suggests some effects might be smaller (3, 4). Especially given that effects of OCD on cardiometabolic traits are hypothesized to be relatively distal, greater power could be required to detect them.

The negative genetic correlation between OCD and BMI, although consistent with previous studies (7, 13), is particularly intriguing given that the observed association at the population level is positive (3). Similarly, negative genetic correlations have been described between BMI and schizophrenia (13, 32, 33), even when obesity is observed to be up to four times more common in individuals with schizophrenia than in the general population (34).

One possible explanation could be that diagnosed OCD and sub-threshold OCD traits have different directions of association with BMI. In clinical OCD populations, there is a much higher prevalence of obesity, which might be (to some degree) a result of medication-related weight gain (35). However, the GWAS of BMI will only contain population prevalence OCD cases (1-2%) (1), with perhaps even fewer individuals taking OCD medication. Therefore, if the observed positive association between having OCD and higher BMI is due to the consequence of getting a diagnosis and subsequently receiving medication, then this might not be observed in population GWAS of cardiometabolic phenotypes (and consequently in genetic correlations). This might explain why genetic correlations with BMI are not in the expected direction for relatively rare disorders (e.g., schizophrenia and OCD), but are in the expected positive direction for more prevalent disorders (e.g., depression) (36). Alternatively, the unexpected association could be due to sample selection, with highly selected samples such as 23andMe and UK Biobank contributing to each of these GWAS respectively. These samples are more highly educated, have a higher socioeconomic status, and tend to be healthier, less likely to be overweight, nor have serious mental health conditions (37). If both having OCD and being overweight make people less likely to participate in these samples, this results in collider bias, which could induce a spurious negative association between the two traits, highlighting the need for more representative and diverse samples, particularly within genetic research.

Given the lack of evidence for bi-directional causal effects between OCD and cardiometabolic phenotypes, we did not run the multivariable MR analyses to explore possible mediation through health behaviors such as smoking, unhealthy diet, lack of physical activity, and alcohol consumption. Previous family-based analyses have suggested that these factors may play an important role in the observed associations, as lifestyle behaviors are influenced by non-shared or unique environments. However, these behaviors also have a substantial genetic component (e.g., 38), which we would expect to pick up in GWAS of OCD and cardiometabolic traits, and subsequently result in a genetic correlation. Furthermore, if these behaviors were on the causal pathway from OCD to cardiometabolic outcomes, we would have expected to observe causal effects in the MR analyses. Given that this is not the case, we think it more likely that results are biased by low statistical power, and future, better powered GWAS of OCD might detect both genetic correlations and causal effects in MR analyses as has been the case for different psychiatric disorders (e.g., depression, anxiety, post-traumatic stress disorder) where greater SNP-heritability has been detected (11, 39, 40).

Regardless of the mechanisms, individuals with OCD present with an increased risk of morbidity and mortality due to cardiometabolic conditions. Hence, monitoring of their health status and secondary prevention interventions may need to be included in their treatment plans, and the environmental factors that may contribute to the association can be targeted. Lifestyle interventions have shown to be effective at reducing cardiometabolic-related outcomes in different groups of psychiatric disorders (41), which could also be used in OCD.

### Strengths and limitations

This study is the first one to conduct bi-directional MR analyses to systematically evaluate causal associations between OCD and a broad range of cardiometabolic disorders and traits. A handful of previous studies had only focused on one single phenotype, namely obesity (12, 13) and CVD (11). Additionally, we used the largest OCD GWAS to date (7), which identified new OCD genetic risk loci and multiple credible candidate causal genes. For the cardiometabolic phenotypes, we carefully selected publicly available GWAS data to minimize sample overlap with OCD and reduce bias. Finally, to ensure that our OCD genetic instruments were valid, we used a positive control outcome (i.e., suicide attempts) for which, based on previous studies, we hypothesized a positive genetic correlation and a causal association (15-17).

The results also need to be interpreted in light of some limitations. First, the OCD GWAS had limited power and, when OCD was the exposure in the MR analyses, we only had power to detect medium effect sizes. Therefore, a lack of evidence for causality in the MR analyses should be interpreted with caution, suggesting an absence of evidence rather than evidence of absence. Second, in order to reduce bias from population stratification and to ensure exposure and outcome GWAS summary statistics were from the same underlying population, we restricted to samples from European ancestry populations. As a result, findings may not generalize to other populations. Third, there is known selection bias in several of the largest samples contributing to the GWAS. For example, the UK Biobank sample is highly selected with less than 5% of participants consenting to take part, and those participating being more highly educated and overall healthier than the general UK population (42). If both liability for OCD and liability for cardiometabolic outcomes are negatively associated with participation, then this could induce collider bias, perhaps explaining negative genetic correlations observed. Finally, MR analyses are likely biased by directional horizontal pleiotropy which likely biases estimates away from the null. Given that null effects were observed here, we think that large bias from horizontal pleiotropy is unlikely, especially given that results are relatively consistent across the range of sensitivity methods employed (e.g., MR Egger) which each make different assumptions about the nature of pleiotropy.

## Conclusions

Our findings align with previous literature using complementary methods and indicate that the association between OCD and cardiometabolic conditions is not likely to be the result of shared genetic liability. Further, we found a lack of evidence for causal effects. However, analyses should be interpreted in light of low statistical power especially in the direction of OCD to cardiometabolic outcomes and should be replicated when larger GWAS of OCD are available.

## Data Availability

All data used in the manuscript are publicly available, with the exception of the summary statistics for OCD, which will be made available upon publication of the latest GWAS manuscript.

## Role of the funder/sponsor

The funders had no role in the design and conduct of the study; collection, management, analysis, and interpretation of the data; and preparation, review, or approval of the manuscript; and decisions to submit the manuscript for publication.

* The PGC OCD Working Group includes: Strom, Nora I.; Gerring, Zachary F.; Galimberti, Marco; Yu, Dongmei; Halvorsen, Matthew W.; Abdellaoui, Abdel; Rodriguez-Fontenla, Cristina; Sealock, Julia M.; Bigdeli, Tim; Coleman, Jonathan R. I.; Mahjani, Behrang; Thorp, Jackson, G.; Bey, Katharina Burton, Christie L.; Luykx, Jurjen J.; Zai, Gwyneth; Alemany, Silvia; Andre, Christine; Askland, Kathleen D.; Banaj, Nerisa; Barlassina, Cristina; Becker Nissen, Judith; Bienvenu, O. Joseph; Black, Donald; Bloch, Michael H.; Bäckmann, Julia; Børte, Sigrid; Bosch, Rosa; Breen, Michael; Brennan, Brian P.; Brentani, Helena; Buxbaum, Joseph D.; Bybjerg-Grauholm, Jonas; Byrne, Enda M.; Cabana-Dominguez, Judit; Camarena, Beatriz; Camarena, Adrian; Cappi, Carolina; Carracedo, Angel; Casas, Miguel; Cavallini, Maria Cristina; Ciullo, Valentina; Cook, Edwin H.; Crosby, Jesse; Cullen, Bernadette A.; De Schipper, Elles J.; Delorme, Richard; Djurovic, Srdjan; Elias, Jason A.; Estivill, Xavier; Falkenstein, Martha J.; Fundin, Bengt T.; Garner, Lauryn; German, Chris; Gironda, Christina; Goes, Fernando S.; Grados, Marco A.; Grove, Jakob; Guo, Wei; Haavik, Jan; Hagen, Kristen; Harrington, Kelly; Havdahl, Alexandra; Höffler, Kira D.; Hounie, Ana G.; Hucks, Donald; Hultman, Christina; Janecka, Magdalena; Jenike, Eric; Karlsson, Elinor K.; Kelley, Kara; Klawohn, Julia; Krasnow, Janice E.; Krebs, Kristi; Lange, Christoph; Lanzagorta, Nuria; Levey, Daniel; Lindblad-Toh, Kerstin; Macciardi, Fabio; Maher, Brion; Mathes, Brittany; McArthur, Evonne; McGregor, Nathaniel; McLaughlin, Nicole C.; Meier, Sandra; Miguel, Euripedes C.; Mulhern, Maureen; Nestadt, Paul S.; Nurmi, Erika L.; O’Connell, Kevin S.; Osiecki, Lisa; Ousdal, Olga Therese; Palviainen, Teemu; Pedersen, Nancy L.; Piras, Fabrizio; Piras, Federica; Potluri, Sriramya; Rabionet, Raquel; Ramirez, Alfredo; Rauch, Scott; Reichenberg, Abraham; Riddle, Mark A.; Ripke, Stephan; Rosário, Maria C.; Sampaio, Aline S.; Schiele, Miriam A.; Skogholt, Anne Heidi; Sloofman, Laura G.; Smit, Jan; Soler Artigas, María; Thomas, Laurent F.; Tifft, Eric; Vallada, Homero; van Kirk, Nathanial; VeenstraVanderWeele, Jeremy; Vulink, Nienke N.; Walker, Christopher P.; Wang, Ying; Wendland, Jens R.; Winsvold, Bendik S.; Yao, Yin; Zhou, Hang; 23andMe Research Team; VA Million Veteran Program; Estonian Biobank; CoGa research team; iPSYCH; HUNT research team; NORDiC research team; Agrawal, Arpana; Alonso, Pino; Berberich, Götz; Bucholz, Kathleen K.; Bulik, Cynthia M.; Cath, Danielle; Denys, Damiaan; Eapen, Valsamma; Edenberg, Howard; Falkai, Peter; Fernandez, Thomas V.; Fyer, Abby J.; Gaziano, J. M.; Geller, Dan A.; Grabe, Hans J.; Greenberg, Benjamin D.; Hanna, Gregory L.; Hickie, Ian B.; Hougaard, David M.; Kathmann, Norbert; Kennedy, James; Lai, Dongbing; Landén, Mikael; Le Hellard, Stéphanie; Leboyer, Marion; Lochner, Christine; McCracken, James T.; Medland, Sarah E.; Mortensen, Preben B.; Neale, Benjamin M.; Nicolini, Humberto; Nordentoft, Merete; Pato, Michele; Pato, Carlos; Pauls, David L.; Piacentini, John; Pittenger, Christopher; Posthuma, Danielle; Ramos-Quiroga, Josep Antoni; Rasmussen, Steven A.; Richter, Margaret A.; Rosenberg, David R.; Ruhrmann, Stephan; Samuels, Jack F.; Sandin, Sven; Sandor, Paul; Spalletta, Gianfranco; Stein, Dan J.; Stewart, S. Evelyn; Storch, Eric A.; Stranger, Barbara E.; Turiel, Maurizio; Werge, Thomas; Andreassen, Ole A.; Børglum, Anders D.; Walitza, Susanne; Hveem, Kristian; Hansen, Bjarne K.; Rück, Christian; Martin, Nicholas G.; Milani, Lili; Mors, Ole; Reichborn-Kjennerud, Ted; Ribasés, Marta; Kvale, Gerd; Mataix-Cols, David; Domschke, Katharina; Grünblatt, Edna; Wagner, Michael; Zwart, John-Anker; Breen, Gerome; Nestadt, Gerald; Kaprio, Jaakko; Arnold, Paul D.; Grice, Dorothy E.; Knowles, James A.; Ask, Helga; Verweij, Karin J.; Davis, Lea K.; Smit, Dirk J.; Crowley, James J.; Scharf, Jeremiah M.; Stein, Murray B.; Gelernter, Joel; Mathews, Carol A.; Derks, Eske M.; Mattheisen, Manuel

## REFERENCES

1. Ruscio AM, Stein DJ, Chiu WT, Kessler RC. The epidemiology of obsessive-compulsive disorder in the National Comorbidity Survey Replication. Mol Psychiatry. 2010; 15(1):53–63.

2. Fernández de la Cruz L, Isomura K, Lichtenstein P, Larsson H, Kuja-Halkola R, Chang Z, et al. All cause and cause specific mortality in obsessive-compulsive disorder: nationwide matched cohort and sibling cohort study. BMJ. 2024; 384:e077564.

3. Isomura K, Brander G, Chang Z, Kuja-Halkola R, Rück C, Hellner C, et al. Metabolic and cardiovascular complications in obsessive-compulsive disorder: A total population, sibling comparison study with long-term follow-up. Biol Psychiatry. 2018; 84(5):324–31.

4. Isomura K, Sidorchuk A, Brander G, Jernberg T, Rück A, Song H, et al. Risk of specific cardiovascular diseases in obsessive-compulsive disorder. J Psychiatric Res. 2021; 135:189–96.

5. Chen MH, Tsai SJ, Su TP, Li CT, Lin WC, Chen TJ, et al. Increased risk of stroke in patients with obsessive-compulsive disorder: A nationwide longitudinal study. Stroke. 2021; 52(8):2601–8.

6. Holmberg A, Pol-Fuster J, Kuja-Halkola R, Larsson H, Lichtenstein P, Chang Z, et al. Multigenerational family coaggregation study of obsessive-compulsive disorder and cardiometabolic disorders. BMJ Ment Health. 2025; 28(1).

7. Strom NI, Gerring ZF, Galimberti M, Yu D, Halvorsen MW, Abdellaoui A, et al. Genome-wide analyses identify 30 loci associated with obsessive-compulsive disorder. Nat Genet. 2025; in press.

8. Frei O, Smeland O, Holland D, Shadrin A, Thompson W, Andreassen O, et al. Bivariate Gaussian mixture model for GWAS summary statistics. Eur Neuropsychopharm. 2019; 29:S898–S9.

9. Wootton RE, Jones HJ, Sallis HM. Mendelian randomisation for psychiatry: How does it work, and what can it tell us? Mol Psychiatry. 2022; 27(1):53–57.

10. Byrne EM, Yang J, Wray NR. Inference in psychiatry via 2-sample Mendelian randomization -From association to causal pathway? JAMA Psychiatry. 2017; 74(12):1191–2.

11. Yu Y, Yang X, Wu J, Hu G, Bai S, Yu R. A Mendelian randomization study of the effect of mental disorders on cardiovascular disease. Front Cardiovasc Med. 2024; 11:1329463.

12. Chen W, Feng J, Jiang S, Guo J, Zhang X, Zhang X, et al. Mendelian randomization analyses identify bidirectional causal relationships of obesity with psychiatric disorders. J Affect Disord. 2023; 339:807–14.

13. Xiao P, Li C, Mi J, Wu J. Evaluating the distinct effects of body mass index at childhood and adulthood on adult major psychiatric disorders. Sci Adv. 2024; 10(37):eadq2452.

14. International Obsessive Compulsive Disorder Foundation Genetics, OCD Collaborative Genetics Assocition Studies. Revealing the complex genetic architecture of obsessive-compulsive disorder using meta-analysis. Mol Psychiatry. 2018; 23(5):1181–8.

15. Sidorchuk A, Kuja-Halkola R, Runeson B, Lichtenstein P, Larsson H, Rück C, et al. Genetic and environmental sources of familial coaggregation of obsessive-compulsive disorder and suicidal behavior: A population-based birth cohort and family study. Mol Psychiatry. 2021; 26(3):974–85.

16. Krebs G, Mataix-Cols D, Rijsdijk F, Ruck C, Lichtenstein P, Lundstrom S, et al. Concurrent and prospective associations of obsessive-compulsive symptoms with suicidality in young adults: A genetically-informative study. J Affect Dis. 2021; 281:422–30.

17. Tyagi H, Bundies G. Resolving the discrepancies of suicide risk in obsessive-compulsive patients: A review of incidence rates and risk factors of suicide and suicide attempts in OCD. BJPsych Open. 2021; 7:S297–S.

18. Docherty AR, Mullins N, Ashley-Koch AE, Qin X, Coleman JRI, Shabalin A, et al. GWAS meta-analysis of suicide attempt: Identification of 12 genome-wide significant loci and implication of genetic risks for specific health factors. Am J Psychiatry. 2023; 180(10):723–38.

19. Burgess S, Davies NM, Thompson SG. Bias due to participant overlap in two-sample Mendelian randomization. Genet Epidemiol. 2016; 40(7):597–608.

20. Nikpay M, Goel A, Won HH, Hall LM, Willenborg C, Kanoni S, et al. A comprehensive 1,000 genomes-based genome-wide association meta-analysis of coronary artery disease. Nat Genet. 2015; 47(10):1121–30.

21. Kurki MI, Karjalainen J, Palta P, Sipila TP, Kristiansson K, Donner KM, et al. FinnGen provides genetic insights from a well-phenotyped isolated population. Nature. 2023; 613(7944):508–18.

22. Bulik-Sullivan B, Finucane HK, Anttila V, Gusev A, Day FR, Loh PR, et al. An atlas of genetic correlations across human diseases and traits. Nat Genet. 2015; 47(11):1236–41.

23. Sanderson E, Glymour MM, Holmes MV, Kang H, Morrison J, Munafo MR, et al. Mendelian randomization. Nat Rev Methods Primers. 2022; 2.

24. Brion MJ, Shakhbazov K, Visscher PM. Calculating statistical power in Mendelian randomization studies. Int J Epidemiol. 2013; 42(5):1497–501.

25. Sanderson E, Davey Smith G, Windmeijer F, Bowden J. An examination of multivariable Mendelian randomization in the single-sample and two-sample summary data settings. Int J Epidemiol. 2019; 48(3):713–27.

26. Liu M, Jiang Y, Wedow R, Li Y, Brazel DM, Chen F, et al. Association studies of up to 1.2 million individuals yield new insights into the genetic etiology of tobacco and alcohol use. Nat Genet. 2019; 51(2):237–44.

27. Wootton RE, Richmond RC, Stuijfzand BG, Lawn RB, Sallis HM, Taylor GMJ, et al. Evidence for causal effects of lifetime smoking on risk for depression and schizophrenia: A Mendelian randomisation study. Psychol Med. 2020; 50(14):2435–43.

28. Wang Z, Emmerich A, Pillon NJ, Moore T, Hemerich D, Cornelis MC, et al. Genome-wide association analyses of physical activity and sedentary behavior provide insights into underlying mechanisms and roles in disease prevention. Nat Genet. 2022; 54(9):1332–44.

29. Locke AE, Kahali B, Berndt SI, Justice AE, Pers TH, Day FR, et al. Genetic studies of body mass index yield new insights for obesity biology. Nature. 2015; 518(7538):197–206.

30. Meddens SFW, de Vlaming R, Bowers P, Burik CAP, Linner RK, Lee C, et al. Genomic analysis of diet composition finds novel loci and associations with health and lifestyle. Mol Psychiatry. 2021; 26(6):2056–69.

31. Sanderson E, Spiller W, Bowden J. Testing and correcting for weak and pleiotropic instruments in two-sample multivariable Mendelian randomization. Stat Med. 2021; 40(25):5434–52.

32. Bahrami S, Steen NE, Shadrin A, O’Connell K, Frei O, Bettella F, et al. Shared genetic loci between body mass index and major psychiatric disorders: A genome-wide association study. JAMA Psychiatry. 2020; 77(5):503–12.

33. Aoki R, Saito T, Ninomiya K, Shimasaki A, Ashizawa T, Ito K, et al. Shared genetic components between metabolic syndrome and schizophrenia: Genetic correlation using multipopulation data sets. Psychiatry Clin Neurosci. 2022; 76(8):361–6.

34. Correll CU, Detraux J, De Lepeleire J, De Hert M. Effects of antipsychotics, antidepressants and mood stabilizers on risk for physical diseases in people with schizophrenia, depression and bipolar disorder. World Psychiatry. 2015; 14(2):119–36.

35. Albert U, Aguglia A, Chiarle A, Bogetto F, Maina G. Metabolic syndrome and obsessive-compulsive disorder: a naturalistic Italian study. Gen Hosp Psychiatry. 2013; 35(2):154–9.

36. Howard DM, Adams MJ, Clarke TK, Hafferty JD, Gibson J, Shirali M, et al. Genome-wide meta-analysis of depression identifies 102 independent variants and highlights the importance of the prefrontal brain regions. Nat Neurosci. 2019; 22(3):343–52.

37. Fry A, Littlejohns TJ, Sudlow C, Doherty N, Adamska L, Sprosen T, et al. Comparison of sociodemographic and health-related characteristics of UK biobank participants with those of the general population. Am J Epidemiol. 2017; 186(9):1026–34.

38. Maes HH, Sullivan PF, Bulik CM, Neale MC, Prescott CA, Eaves LJ, et al. A twin study of genetic and environmental influences on tobacco initiation, regular tobacco use and nicotine dependence. Psychol Med. 2004; 34(7):1251–61.

39. Lukas E, Veeneman RR, Smit DJA, Ahluwalia TS, Vermeulen JM, Pathak GA, et al. A genetic exploration of the relationship between posttraumatic stress disorder and cardiovascular diseases. Transl Psychiatry. 2025; 15(1):1.

40. Li GH, Cheung CL, Chung AK, Cheung BM, Wong IC, Fok MLY, et al. Evaluation of bi-directional causal association between depression and cardiovascular diseases: A Mendelian randomization study. Psychol Med. 2022; 52(9):1765–76.

41. Bradley T, Campbell E, Dray J, Bartlem K, Wye P, Hanly G, et al. Systematic review of lifestyle interventions to improve weight, physical activity and diet among people with a mental health condition. Syst Rev. 2022; 11(1):198.

42. Munafo MR, Tilling K, Taylor AE, Evans DM, Davey Smith G. Collider scope: When selection bias can substantially influence observed associations. Int J Epidemiol. 2018; 47(1):226–35.

43. Shah S, Henry A, Roselli C, Lin H, Sveinbjornsson G, Fatemifar G, et al. Genome-wide association and Mendelian randomisation analysis provide insights into the pathogenesis of heart failure. Nat Commun. 2020; 11(1):163.

44. Nolte IM, Munoz ML, Tragante V, Amare AT, Jansen R, Vaez A, et al. Genetic loci associated with heart rate variability and their effects on cardiac disease risk. Nat Commun. 2017; 8:15805.

45. Willer CJ, Schmidt EM, Sengupta S, Peloso GM, Gustafsson S, Kanoni S, et al. Discovery and refinement of loci associated with lipid levels. Nat Genet. 2013; 45(11):1274–83.

46. Morris AP, Voight BF, Teslovich TM, Ferreira T, Segre AV, Steinthorsdottir V, et al. Large-scale association analysis provides insights into the genetic architecture and pathophysiology of type 2 diabetes. Nat Genet. 2012; 44(9):981–90.

47. Malik R, Chauhan G, Traylor M, Sargurupremraj M, Okada Y, Mishra A, et al. Multiancestry genome-wide association study of 520,000 subjects identifies 32 loci associated with stroke and stroke subtypes. Nat Genet. 2018; 50(4):524–37.

48. Wain LV, Vaez A, Jansen R, Joehanes R, van der Most PJ, Erzurumluoglu AM, et al. Novel blood pressure locus and gene discovery using genome-wide association study and expression data sets from blood and the kidney. Hypertension. 2017; 70(3):e4–e19.

49. Thibord F, Klarin D, Brody JA, Chen MH, Levin MG, Chasman DI, et al. Cross-ancestry investigation of venous thromboembolism genomic predictors. Circulation. 2022; 146(16):1225–42.

