## Supplementary for "Are there causal associations between obsessive-compulsive disorder and cardiometabolic phenotypes? A genetic correlation and bi-directional Mendelian randomization study"

**TABLE**

**Table 1.** Genome-wide summary statistics used in genetic correlations and Mendelian randomization analyses.

| **Cardiometabolic phenotype** | **Reference** | **Date** | **N case** | **N control** | **Total N** | **N GWS SNPs** | **Estimated overlap** |
| --- | --- | --- | --- | --- | --- | --- | --- |
| OCD (with 23andMe) | Strom et al. (7) | 2024 | 53,660 | 2,044,417 | 2,098,077 | 29 | - |
| OCD (without 23andMe) | Strom et al. (7) | 2024 | 23,493 | 1,114,613 | 1,138,106 | - | - |
| Coronary artery disease | Nikpay et al. (20) | 2015 | 22,233 | 64,762 | 86,995 | 42 | 0% |
| Myocardial infarction | Nikpay et al. (20) | 2015 | 48,371 | 123,504 | 171,875 | 26 | 0% |
| Heart failure | Shah et al. (43) | 2020 | 47,309 | 930014 | 977,323 | 12 | 18.8% |
| Heart rate variability | Nolte et al. (44) | 2017 | - | - | 53,174 | 6 | 0% |
| Triglycerides | Willer et al. (45) | 2013 | - | - | 188,578 | 54 | 0.03% |
| Total cholesterol | Willer et al. (45) | 2013 | - | - | 188,578 | 87 | 0.03% |
| HDL cholesterol | Willer et al. (45) | 2013 | - | - | 188,578 | 87 | 0.03% |
| LDL cholesterol | Willer et al. (45) | 2013 | - | - | 188,578 | 75 | 0.03% |
| Type 2 diabetes | Morris et al. (46) | 2012 | 12,171 | 56,862 | 69,033 | 33 | 0.12% |
| Body mass index | Locke et al. (29) | 2015 | - | - | 339,224 | 79 | 0.06% |
| Ischemic stroke | Malik et al. (47) | 2018 | 67,162 | 454,450 | 521,612 | 8 | 0% |
| Systolic blood pressure (without UKBB) | Wain et al. (48) | 2017 | - | - | 150,134 | 80 | 0% |
| Diastolic blood pressure (without UKBB) | Wain et al. (48) | 2017 | - | - | 150,134 | 77 | 0% |
| Thrombosis | Thibord et al. (49) | 2022 | 62,879 | 932,985 | 995,864 | 148 | 2.45% |

*Note:* N GWS SNPs = the number of genome-wide significant SNPs for each exposure after restricting to independent variants (kb window = 10,000, r^2^ = 0.001). Estimated overlap = Percentage sample overlap estimated at the maximum possible cohort overlap as a percentage of the Strom et al. (2024) OCD GWAS (with 23andMe).

*Abbreviations:* HDL, high-density lipoprotein; LDL, low-density lipoprotein; OCD, obsessive-compulsive disorder UKBB, United Kingdom Biobank.

**FIGURES**

**Figure 1.** Genetic correlations between OCD and cardiometabolic phenotypes.

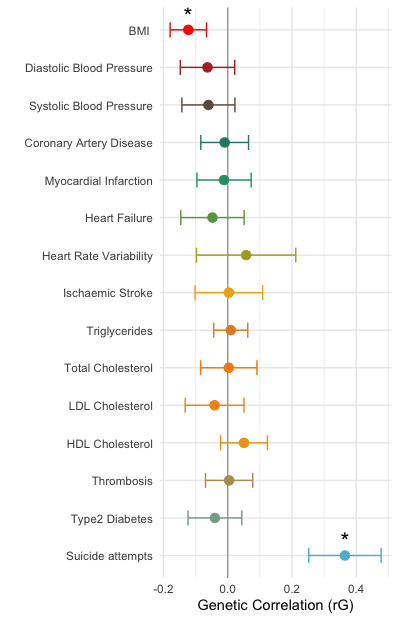

*Abbreviations:* BMI, body mass index; HDL, high-density lipoprotein; LDL, low-density lipoprotein.

**Figure 2.** Univariable Mendelian randomization results (inverse variance weighted) for liability to obsessive-compulsive predicting cardiometabolic phenotypes.
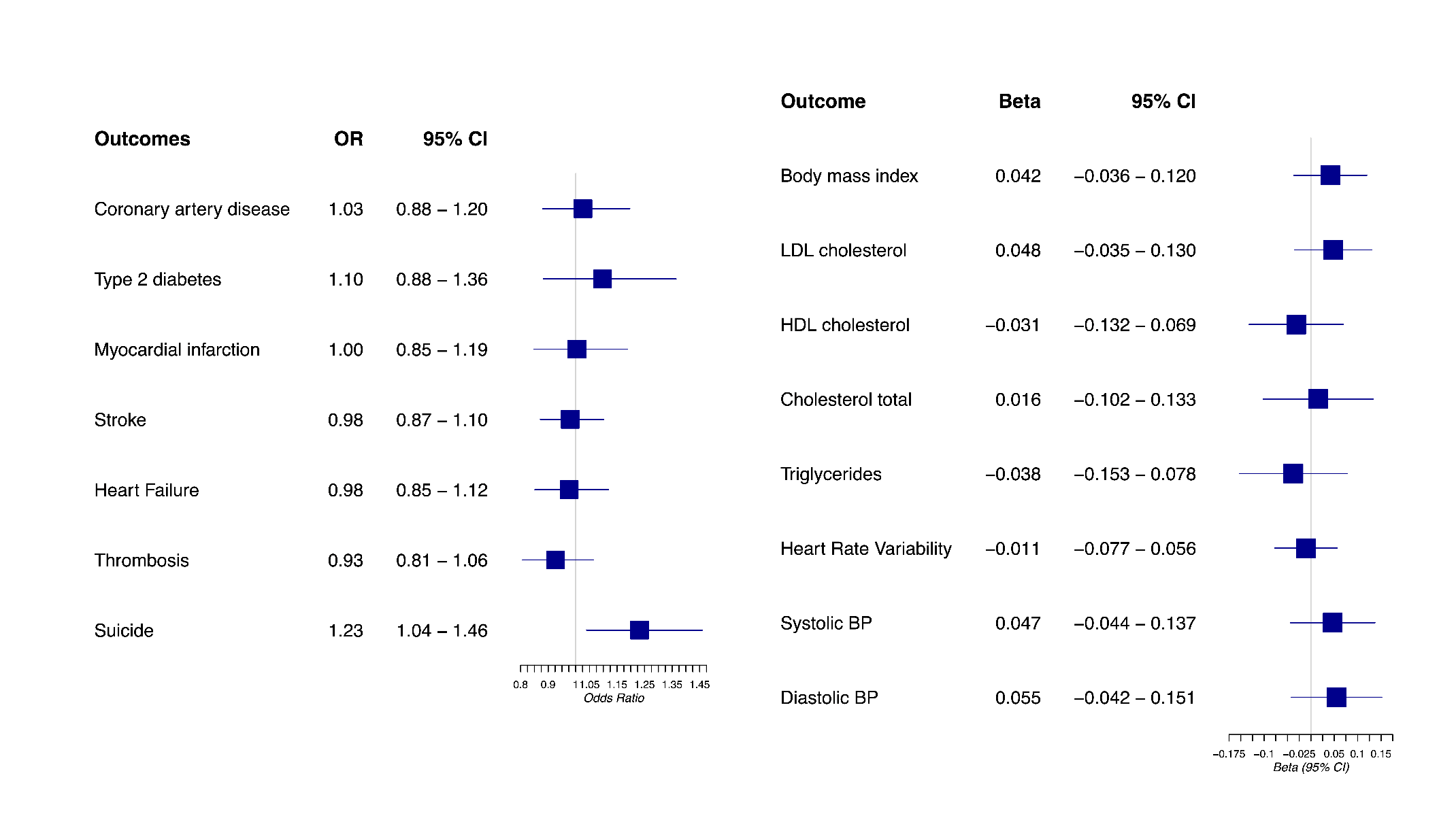
*Abbreviations:* BP, blood pressure; HDL, high-density lipoprotein; LDL, low-density lipoprotein.

**Figure 3.** Univariable Mendelian randomization results (inverse variance weighted) for cardiometabolic phenotypes predicting odds of obsessive-compulsive disorder.

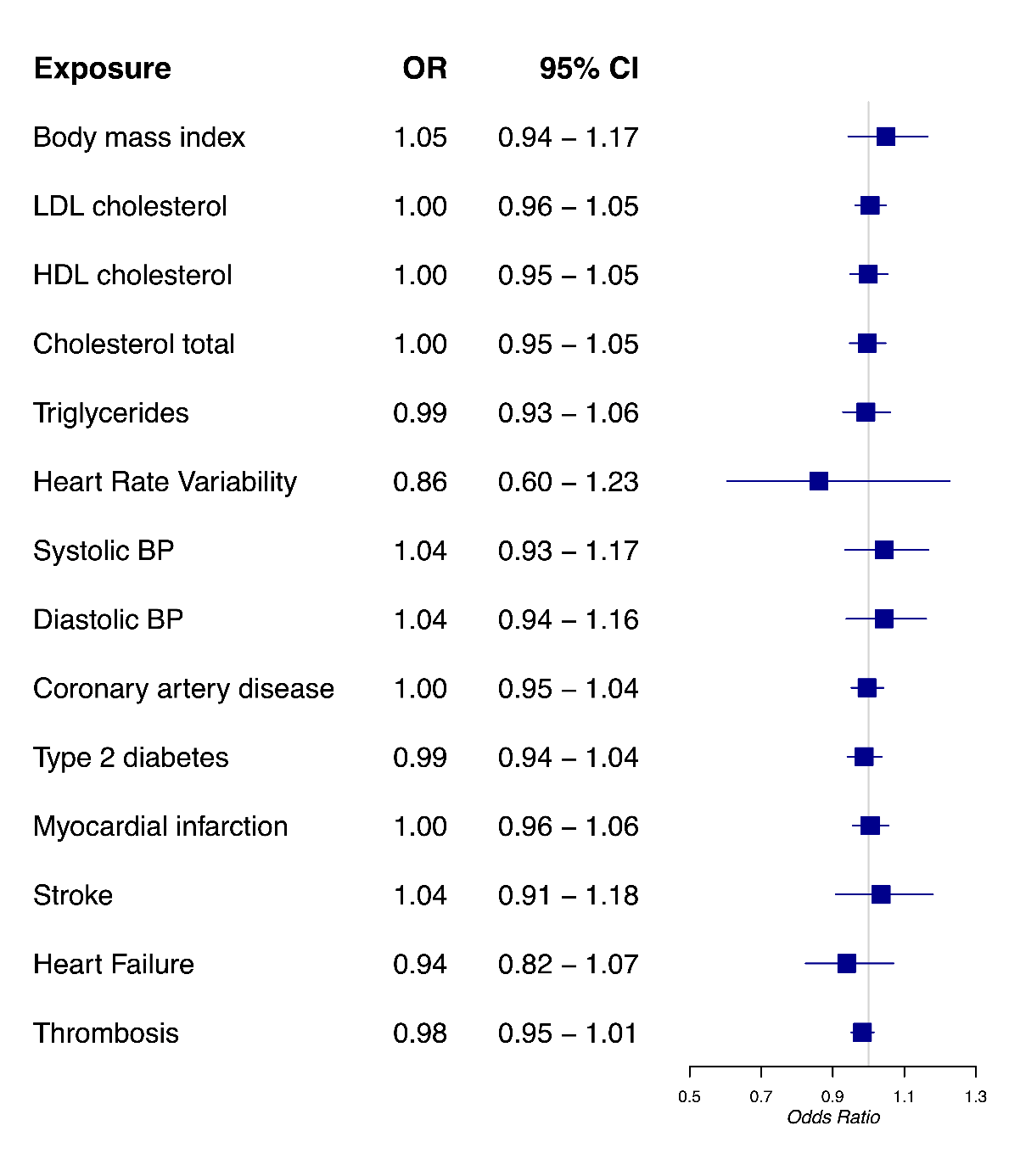

*Abbreviations:* BP, blood pressure; HDL, high-density lipoprotein; LDL, low-density lipoprotein.

**SUPPLEMENTARY MATERIAL**

**Supplementary Table 1.** Power calculations for two-sample Mendelian randomization analysis.

| **Exposure** | **Outcome** | **R^2^** | **Outcome N** | **Outcome proportion cases** | **80% Power OR** | **Effect Size** |
| --- | --- | --- | --- | --- | --- | --- |
| OCD | Coronary artery disease | 0.055% | 86,995 | 0.256 | 2.1 | Medium |
| OCD | Myocardial infarction | 0. 055% | 171,875 | 0.281 | 1.75 | Medium |
| OCD | Heart failure | 0. 055% | 977,323 | 0.048 | 1.57 | Medium |
| OCD | Type 2 diabetes | 0. 055% | 69,033 | 0.176 | 2.35 | Medium |
| **Exposure** | **Outcome** | **R^2^** | **Outcome N** |  | **80% Power Beta** | **Effect Size** |
| OCD | Heart rate variability | 0. 055% | 53,174 | - | 0.46 | Medium |
| OCD | Triglycerides | 0. 055% | 188,578 | - | 0.27 | Medium |
| OCD | Total cholesterol | 0. 055% | 188,578 | - | 0.27 | Medium |
| OCD | HDL cholesterol | 0. 055% | 188,578 | - | 0.27 | Medium |
| OCD | LDL cholesterol | 0. 055% | 188,578 | - | 0.27 | Medium |
| OCD | Body mass index | 0. 055% | 339,224 | - | 0.2 | Medium |
| OCD | Ischemic stroke | 0. 055% | 521,612 | - | 0.163 | Small |
| OCD | Blood pressure | 0. 055% | 150,134 | - | 0.3 | Medium |
| **Exposure** | **Outcome** | **R^2^** | **Outcome N** | **Outcome proportion cases** | **80% Power OR** | **Effect Size** |
| Coronary Artery Disease | OCD | 2.88% | 985,631 | 0.038 | 1.09 | Small |
| Myocardial Infarction | OCD | 0.91% | 985,631 | 0.038 | 1.16 | Small |
| Heart Failure | OCD | 0.05% | 985,631 | 0.038 | 1.67 | Medium |
| Type2 Diabetes | OCD | 2.69% | 985,631 | 0.038 | 1.09 | Small |
| Heart Rate Variability | OCD | 0.95% | 985,631 | 0.038 | 1.16 | Small |
| Triglycerides | OCD | 4.58% | 985,631 | 0.038 | 1.07 | Small |
| Total Cholesterol | OCD | 6.14% | 985,631 | 0.038 | 1.06 | Small |
| HDL Cholesterol | OCD | 6.79% | 985,631 | 0.038 | 1.06 | Small |
| LDL Cholesterol | OCD | 6.98% | 985,631 | 0.038 | 1.06 | Small |
| BMI | OCD | 1.53% | 985,631 | 0.038 | 1.12 | Small |
| Ischaemic Stroke | OCD | 0.06% | 985,631 | 0.038 | 1.60 | Medium |
| Systolic Blood Pressure | OCD | 2.51% | 985,631 | 0.038 | 1.10 | Small |
| Diastolic Blood Pressure | OCD | 2.40% | 985,631 | 0.038 | 1.10 | Small |

**Supplementary Table 2.** Tests of instrument strength and suitability of the MR Egger method.

| **Exposure** | **Outcome** | **mF** | **I^2^_GX_ unweighted** | **I^2^_GX_ weighted** |
| --- | --- | --- | --- | --- |
| OCD | Coronary Artery Disease | 18.6 | 0 | 0 |
| OCD | Type 2 Diabetes | 18.74 | 0 | 0.65 |
| OCD | Myocardial Infarction | 18.60 | 0 | 0 |
| OCD | Stroke | 18.60 | 0 | 0 |
| OCD | Suicide Attempts | 18.46 | 0 | 0 |
| OCD | BMI | 18.80 | 0 | 0 |
| OCD | LDL Cholesterol | 18.80 | 0 | 0 |
| OCD | HDL Cholesterol | 18.80 | 0 | 0 |
| OCD | Total Cholesterol | 18.80 | 0 | 0 |
| OCD | Triglycerides | 18.80 | 0 | 0 |
| OCD | Heart Failure | 18.60 | 0 | 0 |
| OCD | Heart Rate Variability | 18.40 | 0 | 0 |
| OCD | Systolic Blood Pressure | 18.46 | 0 | 0 |
| OCD | Diastolic Blood Pressure | 18.46 | 0 | 0 |
| OCD | Thrombosis | 18.60 | 0 | 0 |
| OCD | Myocardial Infarction (FinnGen) | 18.42 | 0 | 0 |
| Coronary Artery Disease | OCD | 63.87 | 0.91 | 0.91 |
| Type 2 Diabetes | OCD | 69.03 | 0.89 | 0.90 |
| Myocardial Infarction | OCD | 61.78 | 0.9 | 0.90 |
| Stroke | OCD | 41.81 | 0 | 0 |
| BMI | OCD | 65.58 | 0.91 | 0.91 |
| LDL Cholesterol | OCD | 181.54 | 0.99 | 0.99 |
| HDL Cholesterol | OCD | 149.95 | 0.98 | 0.98 |
| Total Cholesterol | OCD | 138.26 | 0.98 | 0.98 |
| Triglycerides | OCD | 160.44 | 0.98 | 0.98 |
| Heart Failure | OCD | 41.08 | 0.83 | 0.20 |
| Heart Rate Variability | OCD | 57.21 | 0.8 | 0.67 |
| Systolic Blood Pressure | OCD | 47.59 | 0.72 | 0.53 |
| Diastolic Blood Pressure | OCD | 45.63 | 0.75 | 0.59 |
| Thrombosis | OCD | 122.86 | 0.98 | 0.98 |
| Myocardial Infarction (FinnGen) | OCD | 67.13 | 0.87 | 0.92 |

**Supplementary Table 3.** Cochran’s Q estimate of heterogeneity in SNP effects.

| **Exposure** | **Outcome** | **Method** | **Q** | **DF** | **P-value** |
| --- | --- | --- | --- | --- | --- |
| OCD | Coronary artery disease | MR Egger | 44.99 | 27 | 0.02 |
|  |  | Inverse variance weighted | 45.9 | 28 | 0.02 |
| OCD | Type 2 diabetes | MR Egger | 29.57 | 23 | 0.16 |
|  |  | Inverse variance weighted | 30.08 | 24 | 0.18 |
| OCD | Myocardial infarction | MR Egger | 44.88 | 27 | 0.02 |
|  |  | Inverse variance weighted | 44.9 | 28 | 0.02 |
| OCD | Stroke | MR Egger | 25.9 | 27 | 0.52 |
|  |  | Inverse variance weighted | 26.11 | 28 | 0.57 |
| OCD | Heart Failure | MR Egger | 52.06 | 27 | 0.003 |
|  |  | Inverse variance weighted | 53.09 | 28 | 0.003 |
| OCD | Thrombosis | MR Egger | 88.1 | 27 | <0.001 |
|  |  | Inverse variance weighted | 94.37 | 28 | <0.001 |
| OCD | Suicide | MR Egger | 43.6 | 25 | 0.01 |
|  |  | Inverse variance weighted | 45.29 | 26 | 0.01 |
| OCD | Myocardial infarction (FinnGen) | MR Egger | 36.94 | 26 | 0.08 |
|  |  | Inverse variance weighted | 36.99 | 27 | 0.1 |
| OCD | Body mass index | MR Egger | 64.71 | 23 | <0.001 |
|  |  | Inverse variance weighted | 64.72 | 24 | <0.001 |
| OCD | LDL cholesterol | MR Egger | 32.33 | 23 | 0.09 |
|  |  | Inverse variance weighted | 38.01 | 24 | 0.03 |
| OCD | HDL cholesterol | MR Egger | 55.13 | 23 | <0.001 |
|  |  | Inverse variance weighted | 66.31 | 24 | <0.001 |
| OCD | Cholesterol total | MR Egger | 54.85 | 23 | <0.001 |
|  |  | Inverse variance weighted | 80.9 | 24 | <0.001 |
| OCD | Triglycerides | MR Egger | 63.08 | 23 | <0.001 |
|  |  | Inverse variance weighted | 91.28 | 24 | <0.001 |
| OCD | Heart Rate Variability | MR Egger | 13.38 | 13 | 0.42 |
|  |  | Inverse variance weighted | 13.47 | 14 | 0.49 |
| OCD | Systolic BP | MR Egger | 84.61 | 25 | <0.001 |
|  |  | Inverse variance weighted | 109.26 | 26 | <0.001 |
| OCD | Diastolic BP | MR Egger | 75.34 | 25 | <0.001 |
|  |  | Inverse variance weighted | 107.08 | 26 | <0.001 |
| Coronary artery disease | OCD | MR Egger | 22.44 | 34 | 0.94 |
|  |  | Inverse variance weighted | 23.14 | 35 | 0.94 |
| Type 2 diabetes | OCD | MR Egger | 16.81 | 22 | 0.77 |
|  |  | Inverse variance weighted | 16.87 | 23 | 0.82 |
| Myocardial infarction | OCD | MR Egger | 10.4 | 22 | 0.98 |
|  |  | Inverse variance weighted | 10.75 | 23 | 0.99 |
| Stroke | OCD | MR Egger | 4.98 | 6 | 0.55 |
|  |  | Inverse variance weighted | 7.18 | 7 | 0.41 |
| Heart Failure | OCD | MR Egger | 6.01 | 8 | 0.65 |
|  |  | Inverse variance weighted | 6.02 | 9 | 0.74 |
| Thrombosis | OCD | MR Egger | 67.41 | 114 | 0.99 |
|  |  | Inverse variance weighted | 67.44 | 115 | 0.99 |
| Myocardial infarction (FinnGen) | OCD | MR Egger | 5.35 | 9 | 0.8 |
|  |  | Inverse variance weighted | 5.55 | 10 | 0.85 |
| Body mass index | OCD | MR Egger | 81.05 | 74 | 0.27 |
|  |  | Inverse variance weighted | 84.23 | 75 | 0.22 |
| LDL cholesterol | OCD | MR Egger | 44.47 | 68 | 0.99 |
|  |  | Inverse variance weighted | 45.31 | 69 | 0.99 |
| HDL cholesterol | OCD | MR Egger | 59.97 | 84 | 0.98 |
|  |  | Inverse variance weighted | 61.5 | 85 | 0.97 |
| Cholesterol total | OCD | MR Egger | 54.26 | 77 | 0.98 |
|  |  | Inverse variance weighted | 54.53 | 78 | 0.98 |
| Triglycerides | OCD | MR Egger | 36.47 | 52 | 0.95 |
|  |  | Inverse variance weighted | 38.65 | 53 | 0.93 |
| Heart Rate Variability | OCD | MR Egger | 0.26 | 3 | 0.97 |
|  |  | Inverse variance weighted | 0.37 | 4 | 0.98 |
| Systolic BP | OCD | MR Egger | 50.93 | 68 | 0.94 |
|  |  | Inverse variance weighted | 51.41 | 69 | 0.94 |
| Diastolic BP | OCD | MR Egger | 53.86 | 67 | 0.88 |
|  |  | Inverse variance weighted | 54.42 | 68 | 0.88 |

**Supplementary Table 4.** MR Egger intercept estimates showing evidence for bias from directional horizontal pleiotropy.

| **Exposure** | **Outcome** | **MR Egger Intercept** | **P-value** |
| --- | --- | --- | --- |
| OCD | Coronary artery disease | -0.010 (-0.048, 0.027) | 0.59 |
| OCD | Type 2 diabetes | 0.017 (-0.036, 0.071) | 0.54 |
| OCD | Myocardial infarction | 0.002 (-0.039, 0.042) | 0.93 |
| OCD | Stroke | 0.007 (-0.022, 0.036) | 0.66 |
| OCD | Heart Failure | 0.012 (-0.02, 0.044) | 0.47 |
| OCD | Thrombosis | 0.022 (-0.009, 0.052) | 0.18 |
| OCD | Suicide | -0.023 (-0.068, 0.023) | 0.33 |
| OCD | Myocardial infarction (FinnGen) | 0.006 (-0.054, 0.066) | 0.85 |
| OCD | Body mass index | 0.001 (-0.018, 0.019) | 0.95 |
| OCD | LDL cholesterol | 0.017 (0.000, 0.034) | 0.06 |
| OCD | HDL cholesterol | 0.022 (0.002, 0.043) | 0.04 |
| OCD | Cholesterol total | 0.036 (0.015, 0.058) | <0.001 |
| OCD | Triglycerides | 0.035 (0.014, 0.057) | <0.001 |
| OCD | Heart Rate Variability | -0.004 (-0.026, 0.019) | 0.77 |
| OCD | Systolic BP | -0.030 (-0.052, -0.008) | 0.01 |
| OCD | Diastolic BP | -0.037 (-0.059, -0.015) | <0.001 |
| Coronary artery disease | OCD | -0.004 (-0.015, 0.006) | 0.41 |
| Type 2 diabetes | OCD | 0.002 (-0.017, 0.021) | 0.81 |
| Myocardial infarction | OCD | -0.004 (-0.017, 0.009) | 0.56 |
| Stroke | OCD | 0.047 (-0.015, 0.11) | 0.19 |
| Heart Failure | OCD | -0.001 (-0.027, 0.024) | 0.92 |
| Thrombosis | OCD | 0.000 (-0.004, 0.005) | 0.87 |
| Myocardial infarction (FinnGen) | OCD | 0.006 (-0.020, 0.033) | 0.67 |
| Body mass index | OCD | 0.006 (-0.001, 0.013) | 0.09 |
| LDL cholesterol | OCD | 0.002 (-0.002, 0.006) | 0.36 |
| HDL cholesterol | OCD | -0.003 (-0.007, 0.002) | 0.22 |
| Cholesterol total | OCD | -0.001 (-0.006, 0.003) | 0.61 |
| Triglycerides | OCD | 0.004 (-0.001, 0.009) | 0.15 |
| Heart Rate Variability | OCD | 0.008 (-0.041, 0.058) | 0.76 |
| Systolic BP | OCD | 0.004 (-0.007, 0.016) | 0.49 |
| Diastolic BP | OCD | 0.004 (-0.007, 0.015) | 0.46 |

**Supplementary Table 5.** Results of Steiger filtering tests to detect possible evidence of reverse causation.

| **Exposure** | **Outcome** | **N False** | **N True** | **% True** |
| --- | --- | --- | --- | --- |
| OCD | Coronary Artery Disease | 1 | 28 | 96.55 |
| OCD | Type 2 Diabetes | 25 | 25 | 50 |
| OCD | Myocardial Infarction | 1 | 28 | 96.55 |
| OCD | Stroke | 0 | 29 | 100 |
| OCD | Heart Failure | 1 | 28 | 96.55 |
| OCD | BMI | 1 | 24 | 96 |
| OCD | LDL Cholesterol | 1 | 24 | 96 |
| OCD | HDL Cholesterol | 1 | 24 | 96 |
| OCD | Total Cholesterol | 2 | 23 | 92 |
| OCD | Heart Rate Variability | 0 | 15 | 100 |
| OCD | Triglycerides | 2 | 23 | 92 |
| OCD | Systolic Blood Pressure | 2 | 25 | 92.59 |
| OCD | Diastolic Blood Pressure | 2 | 25 | 92.59 |
| OCD | Thrombosis | 1 | 28 | 96.55 |
| Coronary Artery Disease | OCD | 0 | 36 | 100 |
| Type 2 Diabetes | OCD | 0 | 24 | 100 |
| Myocardial Infarction | OCD | 0 | 24 | 100 |
| Stroke | OCD | 0 | 8 | 100 |
| Heart Failure | OCD | 0 | 10 | 100 |
| BMI | OCD | 0 | 76 | 100 |
| LDL Cholesterol | OCD | 0 | 70 | 100 |
| HDL Cholesterol | OCD | 0 | 86 | 100 |
| Total Cholesterol | OCD | 0 | 79 | 100 |
| Heart Rate Variability | OCD | 0 | 5 | 100 |
| Triglycerides | OCD | 0 | 54 | 100 |
| Systolic Blood Pressure | OCD | 0 | 70 | 100 |
| Diastolic Blood Pressure | OCD | 0 | 69 | 100 |
| Thrombosis | OCD | 0 | 116 | 100 |

**Supplementary Figure 1.** Full univariable Mendelian randomization results for liability to obsessive-compulsive disorder predicting binary cardiometabolic traits.

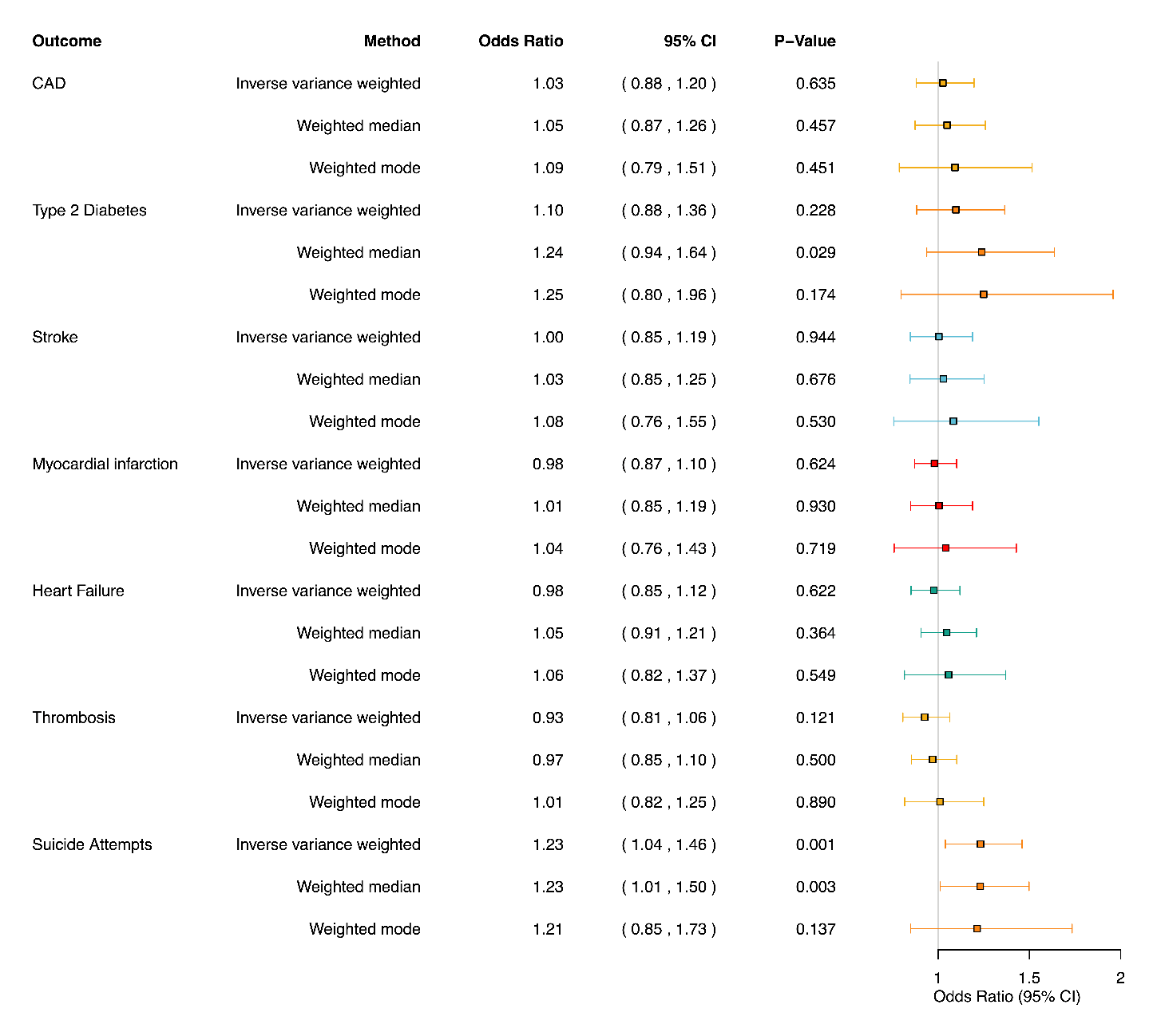

**Supplementary Figure 2.** Full univariable Mendelian randomization results for liability to obsessive-compulsive disorder predicting continuous cardiometabolic traits.

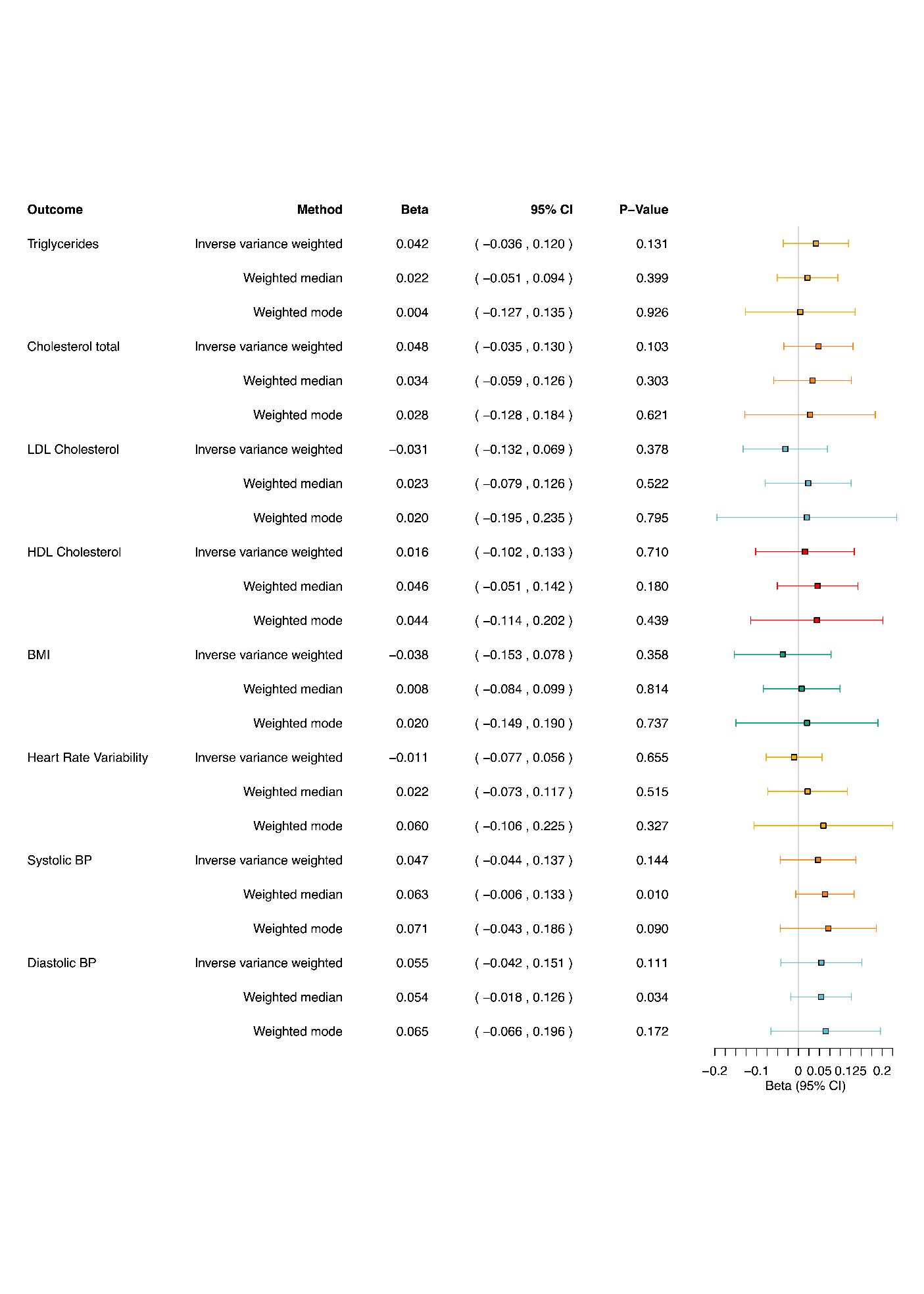

**Supplementary Figure 3a.** Full univariable Mendelian randomization results for cardiometabolic traits predicting liability to obsessive-compulsive disorder.

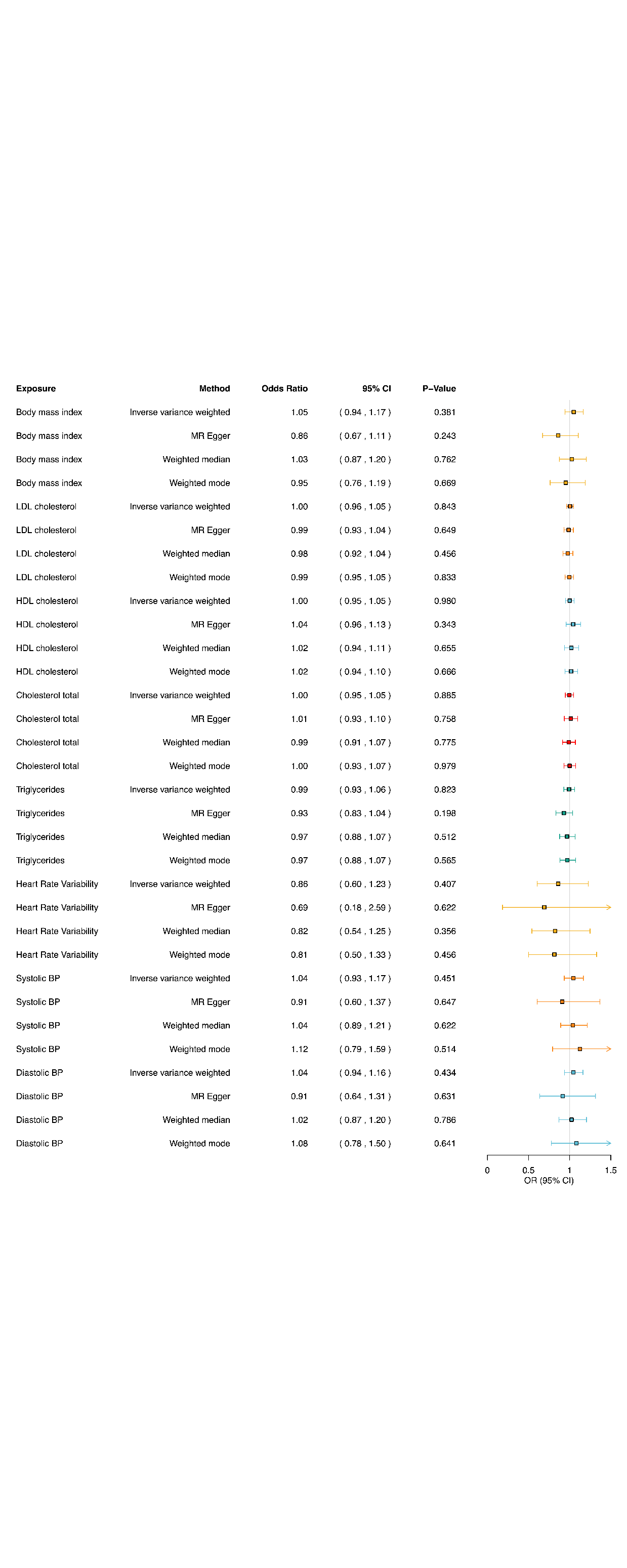

**Supplementary Figure 3b.** Full univariable Mendelian randomization results for cardiometabolic traits predicting liability to obsessive-compulsive disorder.

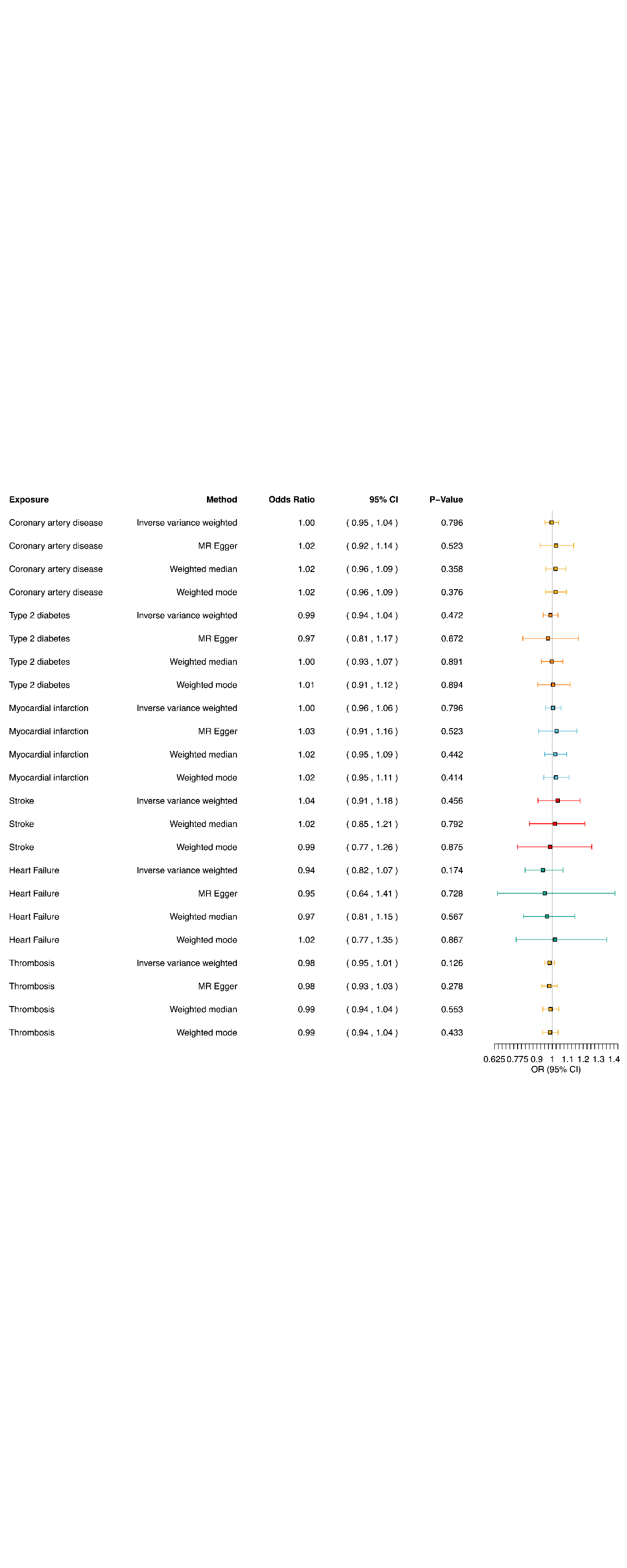

**Supplementary Figure 4.** Scatter plot of SNP-exposure effects on obsessive-compulsive disorder against SNP-outcome effects on suicide attempts.

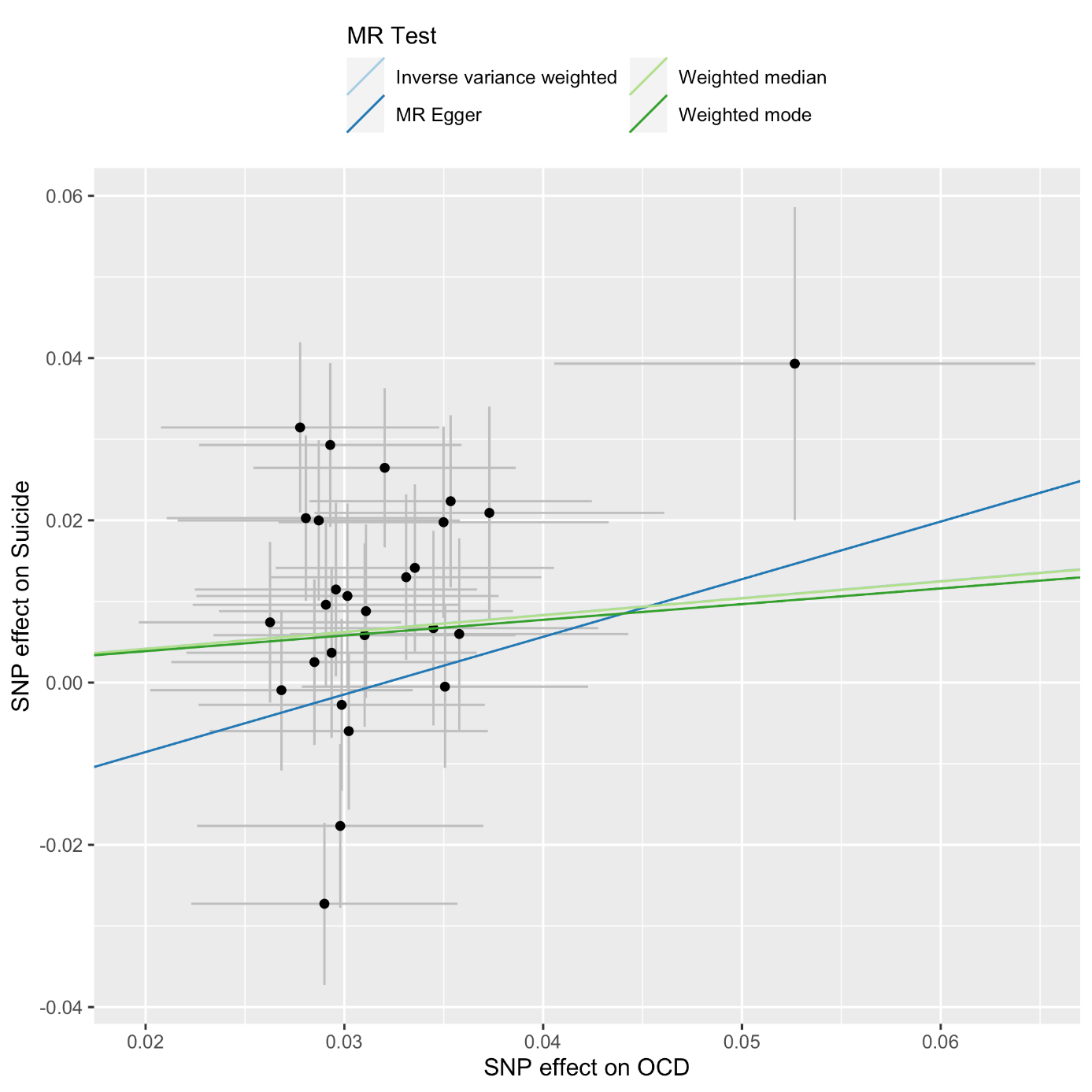

**Supplementary Figure 5.** Leave-one-out SNP plot of obsessive-compulsive disorder on suicide attempts.

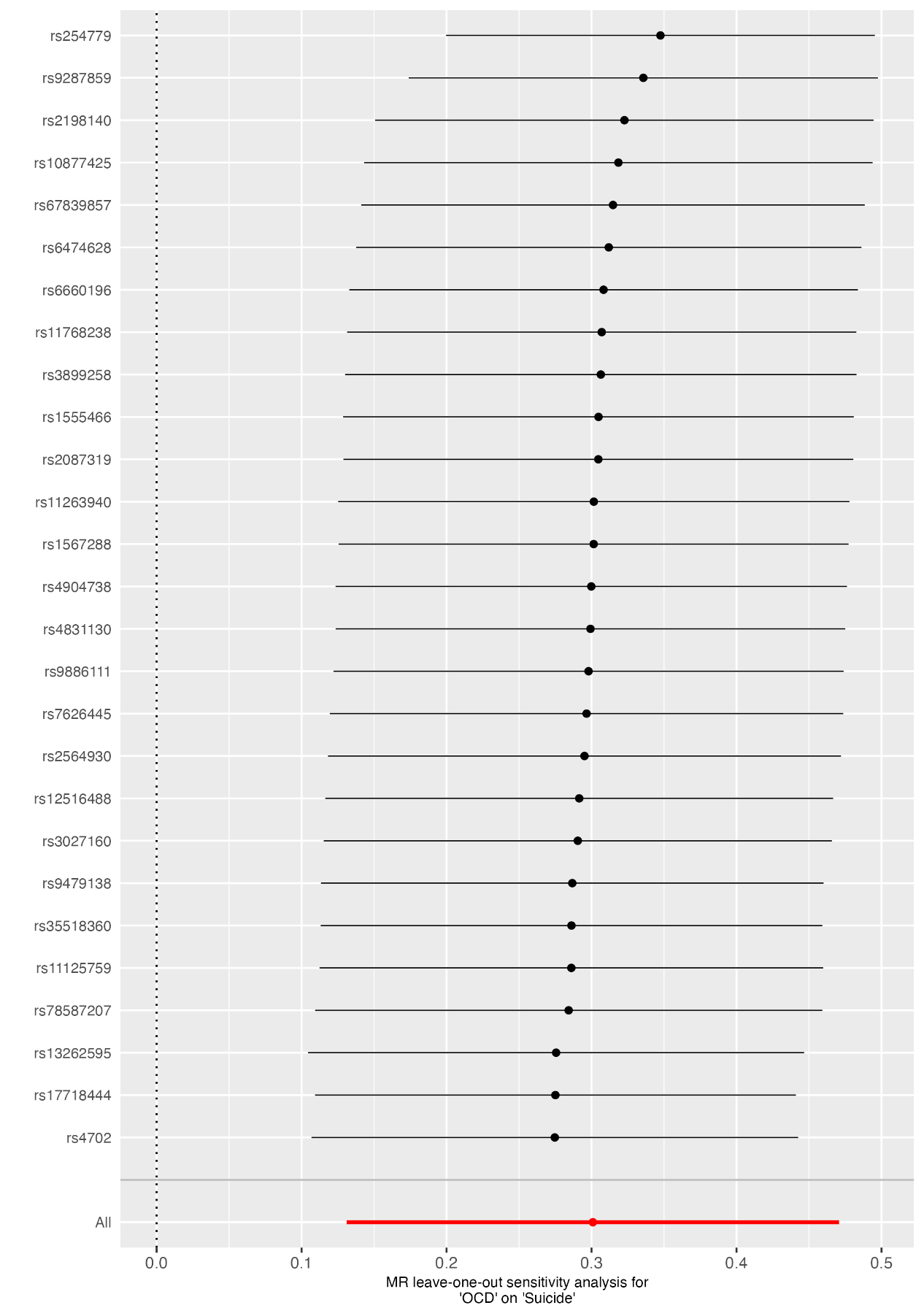

**Supplementary Note S1.** Sensitivity test due to mixed ancestry summary statistics for myocardial infarction.

Nikpay et al.’s (20) GWAS of myocardial infarction (MI) contains >70% European ancestry participants, and the European ancestry only effect sizes were not available. Therefore, we conducted a sensitivity analysis using the myocardial infarction GWAS summary statistics from the FinnGen consortium (21) (details available at: <https://gwas.mrcieu.ac.uk/datasets/finn-b-I9_MI/>).

In the figures below, we plot the results of the Nikpay et al.’s (20) GWAS against the 2021 FinnGen consortium results (21), which show incredibly high concordance. First (Figure S1A), when MI is the exposure, odds of experiencing OCD were highly consistent. Second (Figure S1B), when OCD was the exposure and MI was the outcome, again with highly consistent results.

In conclusion, given the high concordance between results using both sets of summary statistics, we felt confident that results were not substantially biased by differences in ancestry of underlying GWAS populations.

**Figure S1A.**

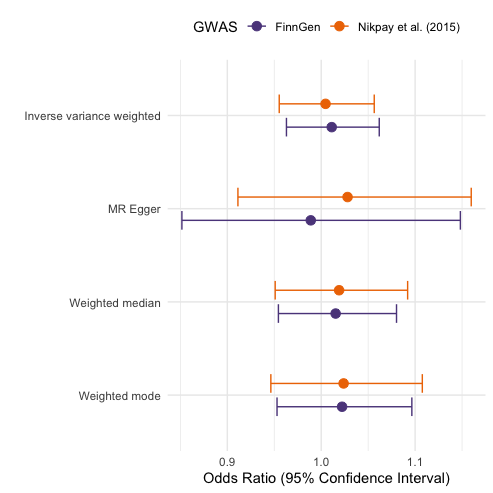

**Figure S1B.**

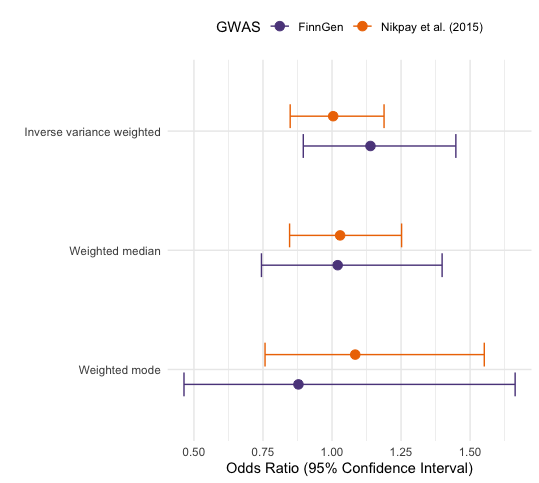
